# Transdiagnostic expression of interoceptive abnormalities in psychiatric conditions

**DOI:** 10.1101/19012393

**Authors:** Hugo D Critchley, Donna L Ewing, Cassandra Gould van Praag, Haniah Habash-Bailey, Jessica A Eccles, Fran Meeten, Sarah N Garfinkel

## Abstract

**Background:** Interoception, the sensing of information about the internal physiological state of the body, is proposed to be fundamental to normal and abnormal affective feelings. We undertook a cross-sectional characterisation of cardiac interoception in patients accessing secondary mental health services to understand how interoceptive abnormalities relate to psychiatric symptoms and diagnoses.

**Methods:** **P**atients attending adult mental health services (205 female, 101 male) and controls (42 female, 21 male) participated. Clinical diagnoses spanned affective disorders, personality disorders and psychoses. Physiological, bio-behavioural and subjective interoceptive measures included: 1) Basal heart rate and heart rate variability (HRV); 2) cardiac afferent effects on emotional processing (cardiac cycle modulation of ratings of fear vs. neutral faces); 3) perceptual accuracy, confidence, and metacognitive insight in heartbeat detection, and; 4) self-reported sensitivity to internal bodily sensations. We tested for transdiagnostic differences between patients and controls, then for correlations between interoceptive measures and affective symptoms, and for group differences across clinical diagnostic categories.

**Results:** Patients differed from controls in HRV, cardiac afferent effects on emotional processing, heartbeat discrimination accuracy, and heartbeat detection confidence. Anxiety and depression symptom severity correlated particularly with self-reported sensitivity to interoceptive experiences. Significant differences between diagnostic categories were observed for HRV, cardiac afferent effects on emotional processing, and subjective interoception. Patients with schizophrenia relative to other diagnoses intriguingly showed opposite cardiac afferent effects on emotion processing.

**Conclusions:** This multilevel characterisation identified interoceptive differences associated with psychiatric symptoms and diagnoses. Interoceptive mechanisms have potential value for the clinical stratification and therapeutic targeting of psychiatric disorders.

## INTRODUCTION

Neurochemical or psychological models dominate understanding of mental illnesses. Available treatments reflect this dichotomy. However, more integrative approaches are emerging with increasing attention to the role of the body in psychological health^1-4^. Interoception refers to the processing and representation of bodily physiology by the central nervous system^1^. Interoception supports physical health by coordinating homeostatic and predictive (allostatic) autonomic and behavioural responses. Psychologically, interoceptive representations underpin associated motivational and emotional feeelings^5-9^. Thus, interoception mediates psychological consequences of poor physical health and moderates the physical impact of negative psychological states (e.g. stress-related cardiovascular morbidity)^3,10-12^.

Within psychiatry, interoception is in the foreground of eating disorders, somatization, and addiction. Yet more broadly, interoceptive feelings underlie mood and anxiety symptoms^1-3^. Moreover, in contemporary models of consciousness, interoception is proposed to be fundamental to self-representation (‘biological self’), supporting coherent continuity of experience^12-15^. Interoceptive dysfunction may thus manifest as disturbances of conscious selfhood e.g. dissociation, depersonalisation, and related psychotic phenomena^4,16-18^. If interoception is central to psychological health, we need to understand its contributions to mental disorders. The Research Domain Criteria initiative (RDoC) of the National Institute of Mental Health (USA)^19^ propose a fresh transdiagnostic biological taxonomy for mental illness towards better treatment targets. RDoC’s major functional domains are *negative valence, positive/reward valence, cognitive systems, systems for social processes* (including self-representation) and *arousal /modulatory systems*^19^. Arguably, interoception is a more fundamental domain supporting basic physiological regulation, motivation, emotional feelings, and self-representation. Here, we tested how interoception relates to diagnosis and affective symptoms in psychiatric patients.

Interoception can be conceptualised along a dimensional framework through measures of; 1) neural signalling of internal physiology; 2) influences on perception and cognition; 3) objective accuracy of interoceptive perception; 3) subjective sensitivity (sensibility) to internal sensations, and; 4) metacognitive interoceptive insight (e.g. awareness of the reliability of one’s perception)^20^. Arguably, emotional experience is grounded upon the quality of interoceptive representations, motivating research into individual differences, with a pragmatic emphasis on cardiac interoception^1,13,17,18, 20-28^. Heart rate variability (HRV) reflects the integration of cardiorespiratory signalling with autonomic cardiovascular control^29,30^. Aortic and carotid arterial baroreceptors fire at cardiac systole (when blood is ejected from the heart) to signal to brainstem the strength of each heartbeat. These signals drive the baroreflex to stabilise cardiac output and blood pressure through heartbeat slowing (parasympathetic activation) and reflex vasodilation (sympathetic inhibition)^31^. In arousal/stress states, baroreflex suppression enables heart rate and blood pressure to rise together^32^, reducing HRV. Beyond the baroreflex, arterial baroreceptor firing affects mental processes: Systole inhibits central processing of pain (and neutral stimuli), yet enhances or preserves perceptual processing of fear and threat^33-35^. These effects are relevant to psychopathology: Systolic enhancement of fear/threat processing predicts heightened anxiety in non-clinical individuals^33^.

People also vary in their capacity to perceive consciously internal sensations; greater sensitivity to interoceptive feelings (measureable using questionnaires or behavioural tasks) may predict stronger emotional experiences. Interoceptive sensitivity is reportedly higher among anxiety and panic patients, but lower in depression^20-22,24^. Interoceptive deficits are also associated with psychotic symptoms in schizophrenia^17,18^. On their own, such findings are heuristic due to psychometric limitations of interoceptive tasks^28^.

One influential framework distinguishes between *subjective* (self-report, questionnaire/confidence), *objective* (e.g. accuracy on interoceptive tasks e.g. heartbeat detection), and *metacognitive* (insight, correspondence between objective and subjective measures) dimensions of interoception^1,20^. Discrepancy between subjective and objective measures of interoception may account for affective symptoms, and is a promising target for intervention^20,25^. Interoceptive abilities also predict intuitive decision-making^23^, a ‘stronger representation of self’^13^, and enhanced impulse control^36^, linking predictive interoceptive representations to self-regulation^4,13-16^.

With converging evidence now connecting psychological symptom expression to aspects of cardiac interoception, there is a need for systematic characterisation in clinical patients^1^. Here we examined a representative group of patients accessing secondary mental health services. We predicted transdiagnostic associations between measures of interoception and affective symptoms (especially anxiety).

## MATERIALS AND METHODS

### Research ethics, governance and study sample

The study was approved by the National Research Ethics Service (13/LO/1866MHRNA), and registered (ISRCTN13588109). Patients, 18+ yrs, accessing services for a recorded psychiatric diagnosis were recruited sequentially from secondary care mental health clinics, or self-referred from advertisement in primary care and community settings. Exclusion criteria included global cognitive impairment, neurological conditions, and alcohol intake on day of testing. Structured diagnostic interviews were not part of the approved protocol. Instead, clinical diagnoses by psychiatrists were confirmed from medical records. Here, 19 patients diagnosed with schizophrenia or paranoid schizophrenia were categorised separately from 26 patients diagnosed with schizoaffective disorder, psychosis with affective features, or unspecified psychosis. An anxiety group, comprising generalised anxiety, social anxiety and panic disorder, was distinguished from posttraumatic stress, and obsessive-compulsive disorders (PTSD, OCD).

Control participants, aged 18-65 yrs, with no mental health diagnosis were recruited through poster advertisements. Exclusion criteria were history of mental or systemic medical illness and medication affecting cardiovascular or cognitive function. Assessments took place in university facilities and hospital clinic rooms.

### Assessment of cardiac physiology and interoceptive dimensions

#### Heart Rate and Heart Rate Variability (HRV)

Pulse-oximetry (Nonin Xpod^®^ 3012LP with soft finger-mount^37^ was used to record heart rate (averaged over six epochs of the heartbeat tracking task, below) and HRV, from root mean square of successive pulse differences; RMSSD^38^ (during the cardiac timing task below, representing longest continuous periods of pulse recording).

#### Cardiac timing effects on fear and neutral face processing

Quantification of cardiac interoceptive influences on emotional processing in non-clinical individuals reveal that ratings of perceived fear intensity are enhanced (or preserved), yet ratings of neutral faces are attenuated, at systole relative to diastole. This differential effect predicts anxiety symptoms^33^. This methodology was adapted for laptop assessments with pulse-oximetry. For 80 randomised trials over two blocks, participants viewed neutral or fearful faces (100 ms), rating each face for emotional intensity (Fig 2A). Stimulus onset was either at ventricular systole (onset 300 ms from estimated ECG R-wave) or late diastole (50ms before ECG R-wave); timing determined from preceding three pulse timings. *Post hoc* evaluation enabled exclusion of outlier trials where onsets exceeded +/-75ms windows from intended systolic or diastolic timings (Fig. 2A).

**Figure 1.**
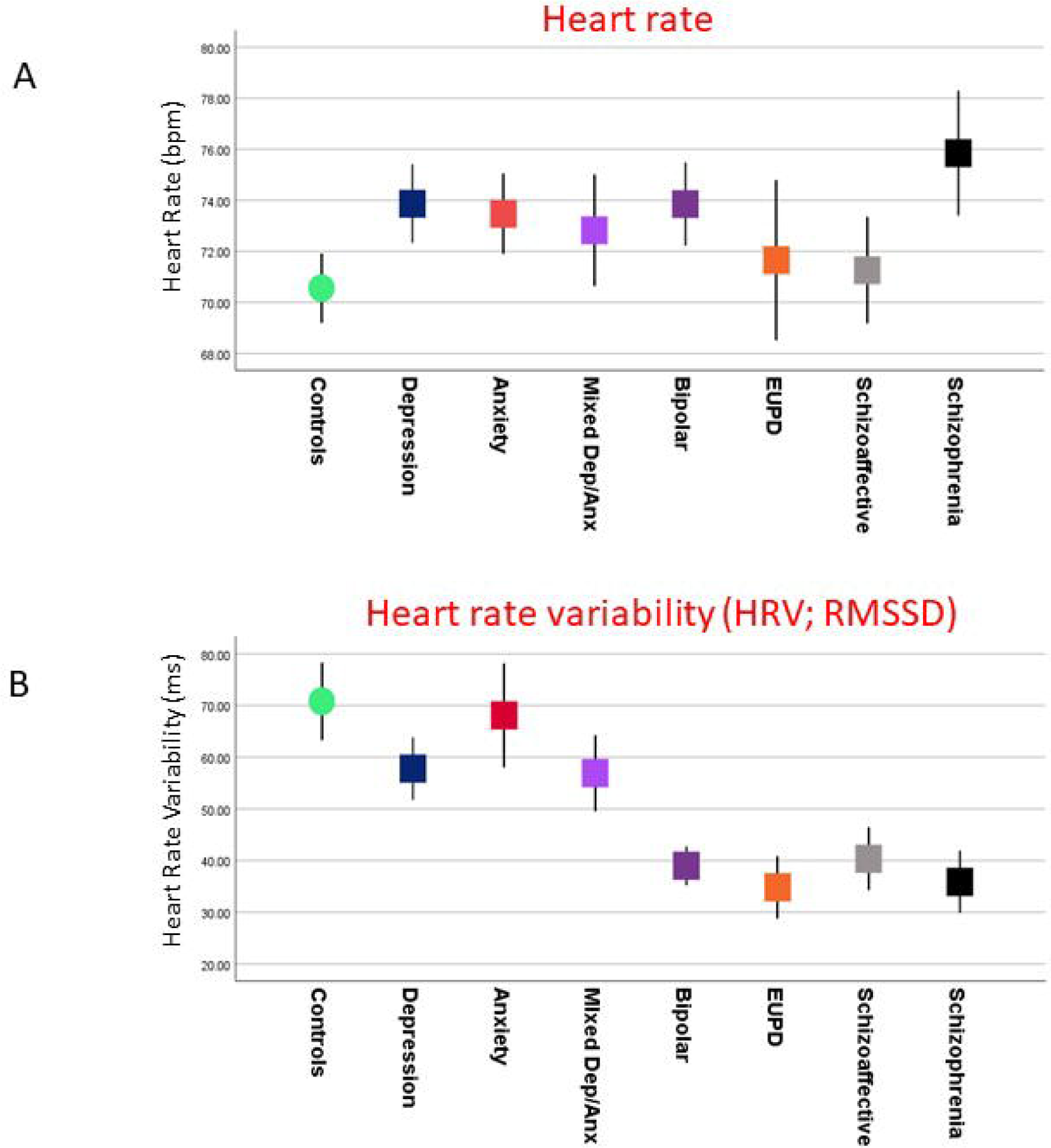
Physiological differences across controls and patients accessing secondary mental health services. **A) Heart rate** Heart rate was recorded for control and patient participants (at rest with interoceptive attentional focus). Differences between controls and the patients as a whole, nor between patient diagnostic subgroups, did not attain threshold significance. Across patients, anxiety symptoms (state) correlated with heart rate (R=0.14 *P*<.05) but not with measures of depression or anxiety. Heart rate did not differentiate patients on or off antipsychotic nor antidepressant medication. **B) Heart rate variability (HRV)** The patient population as a whole manifest lower heart rate variability compared to controls (HRV RMSSD; patients versus controls; ms mean ± SD: 70.19 ± 58.4 vs 51.6 ± 42.6, F(1)=8.8, *P*<.005). Depression symptoms correlated negatively with HRV (RMSSDSD R=-0.15, *P*<.01). There were significant differences in HRV between diagnostic groups of patients (RMSSDSD without age, sex, and BMI covariates: F(6)=3.3, *P*<.01; with covariates included F(6)=2.4, *P*<.05). HRV was no different in people on and off antidepressant medication, but significantly lower in patients on antipsychotic medications (RMSSD; antipsychotic vs no antipsychotic; ms mean ± SD: 42.6 ± 31 vs 59.4 ± 48, F(1)=7.1, *P*<.01).

**Figure 2.**
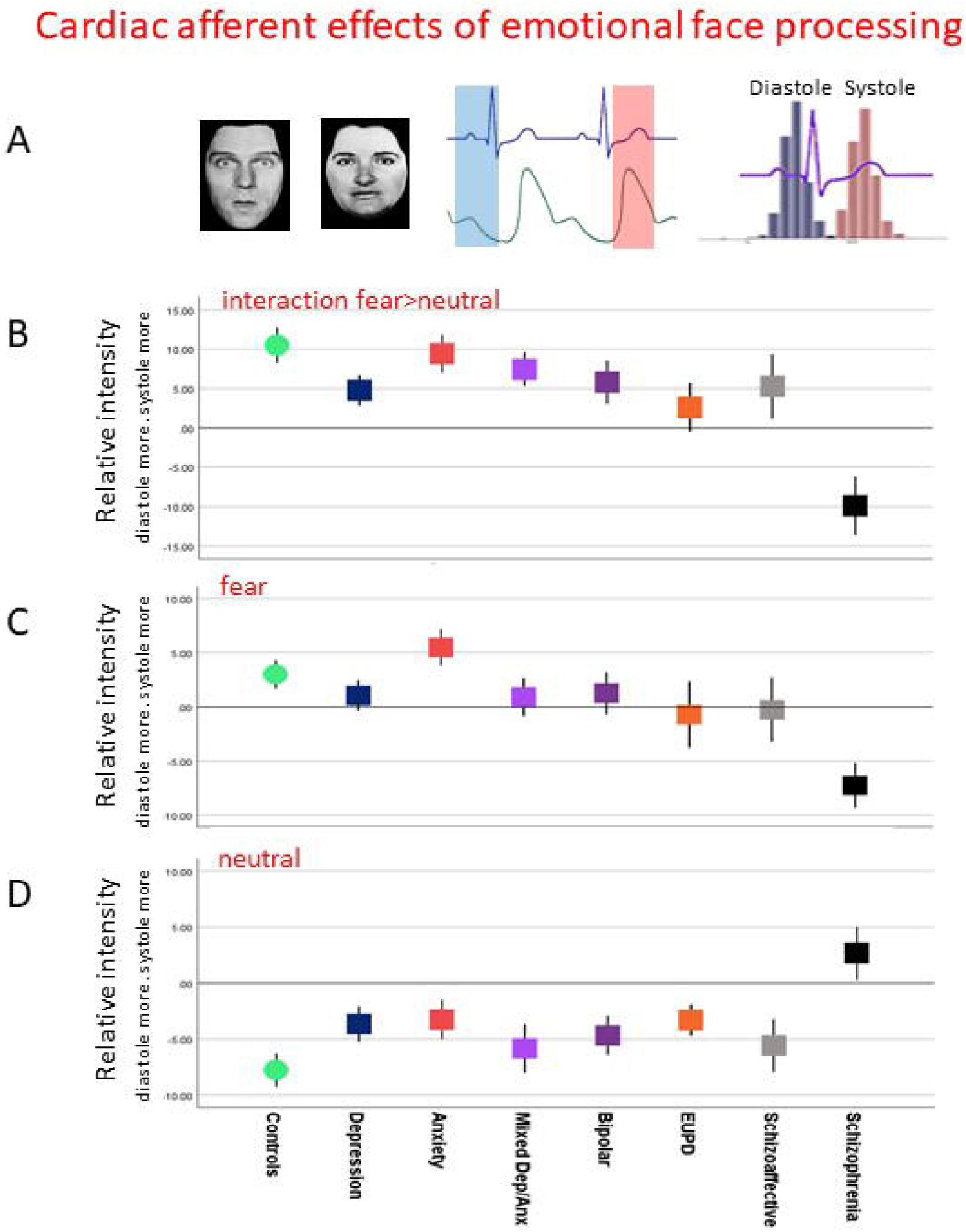
Afferent cardiac (heart timing) effects on processing of fearful and neutral faces. **A) Illustration of heart timing paradigm on emotional processing** Participants were Asked to rate the perceived emotional intensity of fearful face stimuli and neutral face stimuli derived from an emotional face dataset. Stimuli were presented briefly (150ms) to coincide with firing of arterial baroreceptors at cardiac systole (onset ∼250ms from electrocardiograph R wave) or in baroreceptor quiescence at late diastole (onset ∼150ms before - R wave). Timings were determined by a predictive algorithm from earlier heartbeats using pulse oximetry. Post hoc data cleaning excluded trials with onsets outside +/-75ms from intended timings, separating two cardiac timing phases. Previous work demonstrates relative enhancement of ratings for fear vs neutral face expressions at systole vs diastole correlating with reported anxiety symptoms^33^. The figure shows indicative fearful and neutral faces, the onsets of presentation overlaid on ECG and ventricular pressure waveform and the distribution of diastolic and systolic trials. **B) Differences across controls and patients in cardiac afferent effects on emotional face processing: Interaction of fearful vs neutral for ventricular systole vs diastole** Patients, compared to controls, showed a significant difference in cardiac afferent (heart timing; interoceptive baroreceptor signalling at systole) effects on fear vs neutral faces (interaction; patients vs controls; %intensity change mean ± SD: 5.1 ± 16.1 vs 10.5 ± 16.1, F(1,324)=4.8, *P*<.05; F(1,298)=2.0,n.s. with age, sex and BMI covariates). Between patient diagnostic groups, there was a significant differential cardiac timing effect on fear vs neutral face processing (without age, sex, and BMI covariates: F(6,234)=3.1, *P*<.01 with covariates: F(6,234)=2.8, *P*<.05). There was no correlation with affective symptoms nor with antidepressant or antipsychotic medication use. However, a marked difference was observed between schizophrenic patients and other clinical groups, including schizoaffective disorder. **C) Differences across controls and patients in cardiac afferent effects on fearful face processing at ventricular systole vs diastole** Patients, compared to controls, showed no overall difference in cardiac afferent (heart timing; interoceptive baroreceptor signalling at systole) effects on fearful face processing (patients vs controls; %intensity change mean ± SD: 0.7 ± 12.3 vs 3.0 ± 9.8, F(1,331)=1.6, n.s., also n.s. when controlling for age, sex and BMI). Between patient diagnostic groups, there was a trend difference in cardiac timing effects on fearful face processing (without age, sex, and BMI covariates: F(6,238)=2.1, P=0.06 with covariates: F(6,230)=1.7, n.s.). There was no correlation with affective symptoms nor with antidepressant or antipsychotic medication use. However, a significant difference was observed between schizophrenic patients and other clinical groups, including schizoaffective disorder. **D) Differences across controls and patients in cardiac afferent effects on neutral face processing at ventricular systole vs diastole** Patients, compared to controls, showed a significant difference in cardiac afferent (heart timing; interoceptive baroreceptor signalling at systole) effects on neutral face processing (patients vs controls; %intensity change mean ± SD: -4.1 ± 11.5 vs -7.7 ± 10.6, F(1,328)=4.4, *P*<.05, with age, sex and BMI covariates F(1,302)=3.5, P=0.06). Between patient diagnostic groups, there was no significant differential cardiac timing effect on neutral face processing (without age, sex, and BMI covariates: F(6,236)=1.3, n.s., with covariates: F(6,228=1.5, n.s.) There was no correlation with affective symptoms nor with antidepressant or antipsychotic medication use. However, a difference was observed between schizophrenic patients and other clinical groups, including schizoaffective disorder.

#### Heartbeat Tracking Task^27^

Participants counted felt heartbeats at rest, over six randomised 25, 30, 35, 40, 45 and 50 second epochs, then rated their confidence on a visual analogue scale (VAS) from ‘total guess/no heartbeat awareness’ to ‘complete confidence/full perception of heartbeat’. Interoceptive accuracy was quantified by comparing number of reported heartbeats to recorded pulses^20^. Metacognitive insight was computed from accuracy/confidence correspondence (Pearson correlation)^20^.

#### Heartbeat Discrimination Task^26,39,40^

Each trial consisted of 10 auditory tones (440Hz for 100ms) presented either synchronously or delayed relative to heartbeats. Synchronous tones were triggered at the rising edge of the pulse pressure wave. Delayed tones were presented 300ms later. This procedure mirrored established timings of maximum and minimum synchronicity judgements^40,41^. After each trial, participants judged if tones were synchronous or delayed relative to their heartbeats, then rated response confidence. They completed 40 trials over two sessions. Accuracy was measured as proportion of correct trials. Metacognitive insight was computed from accuracy/confidence correspondence (receiver-operating characteristics, ROC, curve)^20^.

#### Porges Body Perception Questionnaire (BPQ)^42,43^ and Interoceptive Trait Prediction Erro

Subjective interoceptive ‘sensibility’ was quantified from self-rating awareness of bodily sensations on the BPQ-awareness scale. Interoceptive Trait Prediction Error (ITPE)^25^ quantified ‘interoceptive surprise’ from discrepancy between subjective s (BPQ-awareness score) and objective (heartbeat tracking accuracy) interoceptive measures^25^.

### Assessment of affective symptoms

Affective symptoms were rated using; 1) *Beck Depression Inventory (BDI)*^44^, a 21 question self-report scale of symptoms associated with depression. 2) *State-Trait Anxiety Inventory (STAI)*^45^, a 40-item self-report questionnaire measuring state (20 items) and trait anxiety (20 items).

### Statistical analyses

Data were analysed parametrically (General Linear Model). Initial analyses compared all patients with controls on physiological and interoceptive measures. Pearson correlations tested for relationships between physiological/interoceptive measures and affective symptoms in patients. Further comparisons tested for effects of antidepressant and antipsychotic medication.

Next, comparisons between diagnostic groups excluded groups with fewer than 10 members and patients with complex mixed diagnoses (excluded: attention deficit hyperactivity disorder N=4; autism N=5; eating disorders N=8; post-traumatic stress disorder N=6; obsessive compulsive disorder N=9; depersonalization disorder N=1; complex mixed diagnoses N=52). Exploratory comparisons of schizophrenia/schizoaffective groups were also undertaken.

To quantify direct relationships with clinical status and symptoms, initially we did not correct for age, sex, or body mass index (BMI). Analyses were then repeated, treating these potential influences on physiology and interoception as confounding covariates to increase inferential precision and balance groups on baseline factors. We did not impute for missing values present across the dataset.

### Supplementary material

Study protocol and full anonymised dataset will be uploaded as Supplementary Information on acceptance of manuscript.

## RESULTS

### Baseline characteristics (Table 1)

Patients were overall older (patients vs controls; yrs mean ± SD: 38.9 ±14.1 vs 35.0 ± 13.2, [F(1,368)=4.1, *P*=.04]) with greater BMI (kg/m^2^: 26.4 ± 7.1 vs 23.0 ± 3.6, [F(1,336)=9.2, *P*=.003]). 56% of patients and 67% of controls were female. Most patients took medication.

**Table 1.** Patient characteristics.

| Group<br>(max N) | Gender | Age<br>yrs | BMI<br>kg/m<sup>2</sup> | HR<br>Bpm | HRV<br>ms | BDI<br>max 63 | STAI-Y1<br>max 80 | STAI-Y2<br>max 80 |
| --- | --- | --- | --- | --- | --- | --- | --- | --- |
| Controls<br>(63) | 42F | 35.0 | 22.97 | 70.55 | 70.82 | 4.87 | 29.88 | 36.92 |
|  | 21M | (13.2) | (3.6) | (8.9) | (58.4) | (6.1) | (7.5) | (8.5) |
| Depression<br>(59) | 34F | 38.4 | 25.35 | 73.87 | 57.78 | 22.34 | 42.17 | 53.92 |
|  | 24M | (15.2) | (7.1) | (11.6) | (45.9) | (13.6) | (13.2) | (12.0) |
| Anxiety<br>(29) | 19F | 35.1 | 23.56 | 73.47 | 68.10 | 17.10 | 42.03 | 55.10 |
|  | 10M | (16.1) | (5.8) | (8.4) | (53.1) | (10.5) | (11.8) | (10.1) |
| Mixed A/D<br>(47) | 32F | 41.5 | 25.25 | 72.83 | 56.90 | 27.78 | 44.40 | 60.87 |
|  | 15M | (12.7) | (5.5) | (14.1) | (46.9) | (13.4) | (11.3) | (8.8) |
| Bipolar<br>(56) | 32F | 42.8 | 27.23 | 73.85 | 38.99 | 22.44 | 38.43 | 53.96 |
|  | 24M | (12.7) | (5.9) | (12.1) | (25.8) | (15.1) | (13.9) | (13.8) |
| EUPD<br>(22) | 15F | 31.5 | 28.91 | 71.65 | 34.82 | 30.59 | 42.91 | 61.36 |
|  | 7M | (13.1) | (11.4) | (13.7) | (27.0) | (16.8) | (13.2) | (11.5) |
| Schizoaffective<br>(25) | 11F | 39.0 | 29.92 | 71.26 | 40.37 | 17.61 | 38.12 | 50.42 |
|  | 13M | (13.4) | (6.1) | (10.0) | (28.5) | (14.6) | (11.4) | (12.6) |
|  | 1NB |  |  |  |  |  |  |  |
| Schizophrenia<br>(19) | 6F | 40.9 | 30.35 | 75.85 | 35.88 | 14.78 | 37.47 | 43.26 |
|  | 12M | (13.7) | (7.2) | (10.1) | (24.6) | (11.3) | (14.8) | (16.3) |
|  | 1NB |  |  |  |  |  |  |  |
| OCD<br>(9) | 4F 7M | 39.0 | 22.78 | 72.61 | 65.96 | 25.67 | 45.00 | 60.44 |
|  |  | (19.4) | (7.2) | (12.0) | (25.9) | (16.0) | (14.3) | (12.6) |
| PTSD<br>(6) | 4F 2M | 39.3 | 28.14 | 71.20 | 45.30 | 35.50 | 46.17 | 63.17 |
|  |  | (11.2) | (5.2) | (10.7) | (29.9) | (13.3) | (4.8) | (11.5) |
| Anorexia<br>(8) | 8F | 29.0 | 19.42 | 69.82 | 72.14 | 18.50 | 42.00 | 53.63 |
|  |  | (8.7) | (2.6) | (9.2) | (43.8) | (12.7) | (9.0) | (11.8) |
| Autism<br>(6) | 4F | 38.83 | 27.95 | 77.28 | 22.10 | 13.67 | 42.33 | 52.00 |
|  | 2M | (19.4) | (7.8) | (8.4) | (8.2) | (5.89) | (8.6) | (11.6) |
| ADHD<br>(4) | 4F | 37.75 | 26.04 | 71.57 | 39.35 | 12.00 | 31.25 | 42.50 |
|  |  | (11.6) | (2.3) | (4.3) | (13.4) | (10.9) | (3.6) | (10.8) |
| Complex<br>(16) | 7F | 38.50 | 25.82 | 69.98 | 80.75 | 15.81 | 38.47 | 56.27 |
|  | 8M | (16.1) | (8.3) | (13.4) | (72.7) | (16.6) | (11.2) | (12.3) |
|  | 1NB |  |  |  |  |  |  |  |

Mean (SD). F=Female, M=Male, NB=non-binary. max N=maximum number in group (not all measure sin all participants – see Supplementary Materials). BMI -= body mass index; HR = heart rate; HRV = heart rate variability; BDI= Beck Depression Inventory; STAI = Spielberger state and Trait anxiety Inventory Y1 = state, Y2 = trait. Anxiety -= generalised anxiety, social anxiety and phobic disorder; Mixed A/D = dual diagnosis of anxiety and depression; EUPD = emotional unstable personality disorder or borderline personality disorder; Schizophrenia = explicit diagnosis of schizophrenia or paranoid schizophrenia diagnosis; Schizoaffective= diagnosis of schizoaffective disorder, affective psychosis or unspecified psychosis; OCD = obsessive compulsive disorder PTSD = post traumatic stress disorder, ADHD attention deficit hyperactivity disorder; Complex =inconclusive, unstated and multiple mixed diagnosis.

### Differences between patient and control populations on cardiac and interoceptive measures

Patients did not differ from controls in heart rate (patients vs controls; bpm 73.0 ± 11.6 vs 70.6 ± 8.9, [F(1,331)=1.9, *P*=.17]), but had lower HRV (RMSSD; patients vs controls; ms: 51.6 ± 42.6 vs 70.8 ± 58.4 [F(1,341)=8.8, *P*=.003]). This difference became non-significant when age, sex, and BMI were included as confounding covariates (Fig. 1, Table 1).

Across all participants (patients plus controls), as anticipated^33^, systole preserved/augmented the rated intensity of fearful faces, and decreased rated intensity of neutral faces, amplifying differences between these emotions (systole versus diastole, interaction i.e. difference between fear vs neutral intensity, 6.0 ± 16.3 [t(324)=6.7, *P*<.001]; fear intensity change 1.1 ± 11.9 [t(331)=1.7, *P*=.09]; neutral intensity change %, - 4.77 ± 11.5 [t(328)=-7.5, *P*<.001]). Inclusion of baseline covariates preserved these effects (interaction F(3,298)=5.15, *P*=.002; fear F(3,303)=1.9, *P*=.14); neutral (F(3,303)=5.4, *P*=.001).

Patients differed from controls in cardiac timing effects on emotional processing (interaction; 5.2 ± 16.1 vs 10.5 ± 16.1, [F(1,324)=4.8, *P*=.03]; fear 0.7 ± 12.3 vs 3.0 ± 9.8, [F(1,331)=1.6, *P*=.20]; neutral -4.1 ± 11.5 vs - 7.7 ± 10.6, [F(1,328)=4.4, *P*=.04]) (Fig. 2; Table 2). Inclusion of baseline covariates attenuated this difference (interaction 5.3 ± 16.3 vs 11.7 ± 16.6, [F(1,298)=2.0, *P*=.16]; fear 2.5 ± 10.6 vs 0.96 ± 12.4, [F(1,303)=0.2, *P*=.66]); neutral -4.1 ± 11.6 vs -9.4 ± 10.6, [F(1,302)=3.5, *P*=.06]) (Fig 2).

**Table 2:**
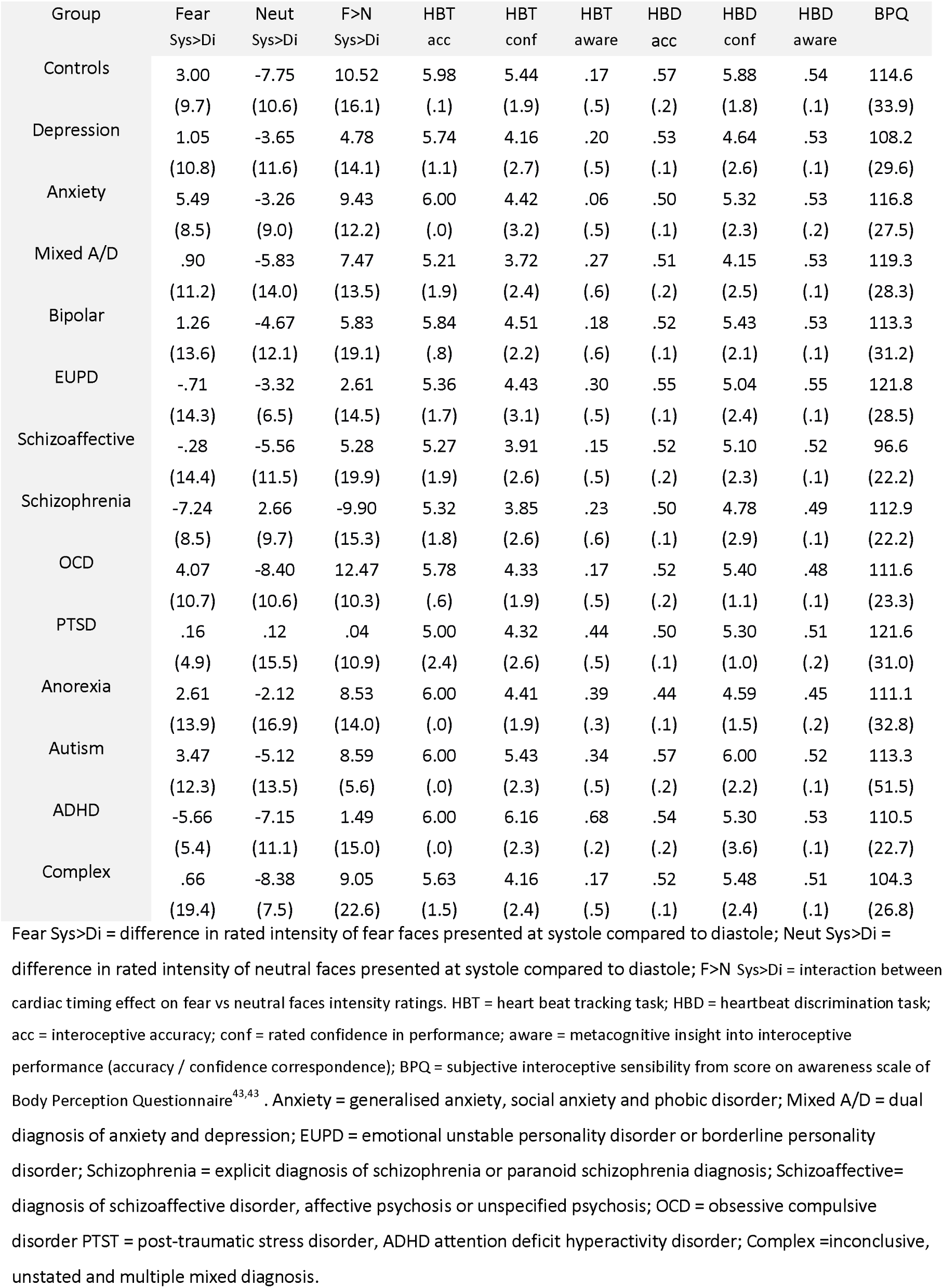
Interoceptive measures across patient groups.

| Group | Fear<br>Sys>Di | Neut<br>Sys>Di | F>N<br>Sys>Di | HBT<br>acc | HBT<br>conf | HBT<br>aware | HBD<br>acc | HBD<br>conf | HBD<br>aware | BPQ |
| --- | --- | --- | --- | --- | --- | --- | --- | --- | --- | --- |
| Controls | 3.00<br>(9.7) | -7.75<br>(10.6) | 10.52<br>(16.1) | 5.98<br>(.1) | 5.44<br>(1.9) | .17<br>(.5) | .57<br>(.2) | 5.88<br>(1.8) | .54<br>(.1) | 114.6<br>(33.9) |
| Depression | 1.05<br>(10.8) | -3.65<br>(11.6) | 4.78<br>(14.1) | 5.74<br>(1.1) | 4.16<br>(2.7) | .20<br>(.5) | .53<br>(.1) | 4.64<br>(2.6) | .53<br>(.1) | 108.2<br>(29.6) |
| Anxiety | 5.49<br>(8.5) | -3.26<br>(9.0) | 9.43<br>(12.2) | 6.00<br>(.0) | 4.42<br>(3.2) | .06<br>(.5) | .50<br>(.1) | 5.32<br>(2.3) | .53<br>(.2) | 116.8<br>(27.5) |
| Mixed A/D | .90<br>(11.2) | -5.83<br>(14.0) | 7.47<br>(13.5) | 5.21<br>(1.9) | 3.72<br>(2.4) | .27<br>(.6) | .51<br>(.2) | 4.15<br>(2.5) | .53<br>(.1) | 119.3<br>(28.3) |
| Bipolar | 1.26<br>(13.6) | -4.67<br>(12.1) | 5.83<br>(19.1) | 5.84<br>(.8) | 4.51<br>(2.2) | .18<br>(.6) | .52<br>(.1) | 5.43<br>(2.1) | .53<br>(.1) | 113.3<br>(31.2) |
| EUPD | -.71<br>(14.3) | -3.32<br>(6.5) | 2.61<br>(14.5) | 5.36<br>(1.7) | 4.43<br>(3.1) | .30<br>(.5) | .55<br>(.1) | 5.04<br>(2.4) | .55<br>(.1) | 121.8<br>(28.5) |
| Schizoaffective | -.28<br>(14.4) | -5.56<br>(11.5) | 5.28<br>(19.9) | 5.27<br>(1.9) | 3.91<br>(2.6) | .15<br>(.5) | .52<br>(.2) | 5.10<br>(2.3) | .52<br>(.1) | 96.6<br>(22.2) |
| Schizophrenia | -7.24<br>(8.5) | 2.66<br>(9.7) | -9.90<br>(15.3) | 5.32<br>(1.8) | 3.85<br>(2.6) | .23<br>(.6) | .50<br>(.1) | 4.78<br>(2.9) | .49<br>(.1) | 112.9<br>(22.2) |
| OCD | 4.07<br>(10.7) | -8.40<br>(10.6) | 12.47<br>(10.3) | 5.78<br>(.6) | 4.33<br>(1.9) | .17<br>(.5) | .52<br>(.2) | 5.40<br>(1.1) | .48<br>(.1) | 111.6<br>(23.3) |
| PTSD | .16<br>(4.9) | .12<br>(15.5) | .04<br>(10.9) | 5.00<br>(2.4) | 4.32<br>(2.6) | .44<br>(.5) | .50<br>(.1) | 5.30<br>(1.0) | .51<br>(.2) | 121.6<br>(31.0) |
| Anorexia | 2.61<br>(13.9) | -2.12<br>(16.9) | 8.53<br>(14.0) | 6.00<br>(.0) | 4.41<br>(1.9) | .39<br>(.3) | .44<br>(.1) | 4.59<br>(1.5) | .45<br>(.2) | 111.1<br>(32.8) |
| Autism | 3.47<br>(12.3) | -5.12<br>(13.5) | 8.59<br>(5.6) | 6.00<br>(.0) | 5.43<br>(2.3) | .34<br>(.5) | .57<br>(.2) | 6.00<br>(2.2) | .52<br>(.1) | 113.3<br>(51.5) |
| ADHD | -5.66<br>(5.4) | -7.15<br>(11.1) | 1.49<br>(15.0) | 6.00<br>(.0) | 6.16<br>(2.3) | .68<br>(.2) | .54<br>(.2) | 5.30<br>(3.6) | .53<br>(.1) | 110.5<br>(22.7) |
| Complex | .66<br>(19.4) | -8.38<br>(7.5) | 9.05<br>(22.6) | 5.63<br>(1.5) | 4.16<br>(2.4) | .17<br>(.5) | .52<br>(.1) | 5.48<br>(2.4) | .51<br>(.1) | 104.3<br>(26.8) |

Fear Sys>Di = difference in rated intensity of fear faces presented at systole compared to diastole; Neut Sys>Di = difference in rated intensity of neutral faces presented at systole compared to diastole; F>N Sys>Di = interaction between cardiac timing effect on fear vs neutral faces intensity ratings. HBT = heart beat tracking task; HBD = heartbeat discrimination task; acc = interoceptive accuracy; conf = rated confidence in performance; aware = metacognitive insight into interoceptive performance (accuracy / confidence correspondence); BPQ = subjective interoceptive sensibility from score on awareness scale of Body Perception Questionnaire<sup>43,43</sup>. Anxiety = generalised anxiety, social anxiety and phobic disorder; Mixed A/D = dual diagnosis of anxiety and depression; EUPD = emotional unstable personality disorder or borderline personality disorder; Schizophrenia = explicit diagnosis of schizophrenia or paranoid schizophrenia diagnosis; Schizoaffective= diagnosis of schizoaffective disorder, affective psychosis or unspecified psychosis; OCD = obsessive compulsive disorder PTST = post-traumatic stress disorder, ADHD attention deficit hyperactivity disorder; Complex =inconclusive, unstated and multiple mixed diagnosis.

On the heartbeat tracking task, patients were significantly less confident than controls (VAS: 4.2 ± 2.6 vs 5.4 ± 1.8; [F(1,333)=8.82, *P*=.003]; with baseline covariates [F(1,318)=10.3, *P*=0.001]). However, there were no group differences in performance accuracy (5.6 ± 1.4 vs 6.0 ± .16, [F(1,348)=3.0, *P*=.09]), nor metacognitive insight (0.21 ± 0.5 vs 0.18 ± 0.5, [F(1,309)=.24, *P*=.62]) (Fig. 3, Table 2).

**Figure 3.**
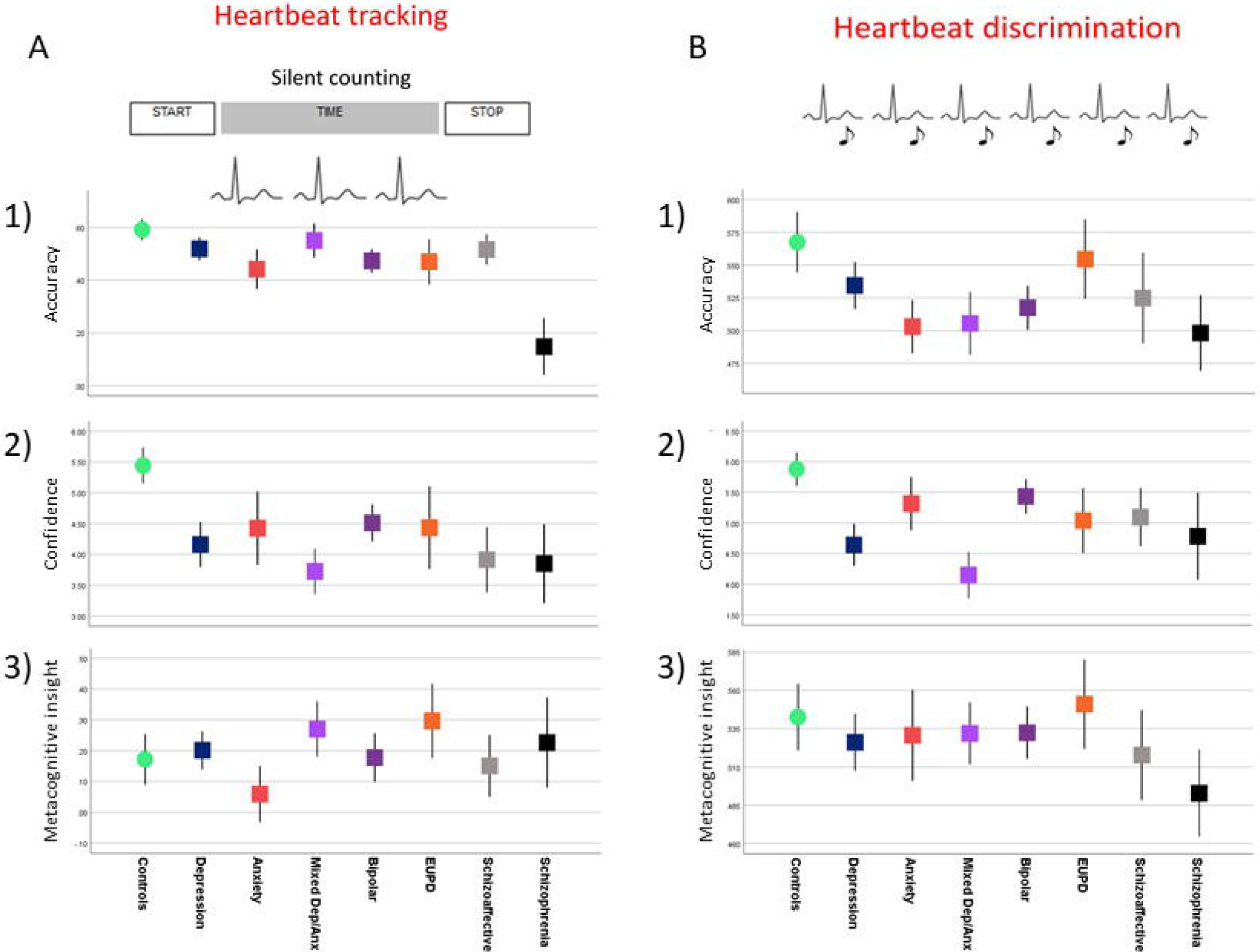
Performance of controls and patients on heartbeat detection tasks. **A) Heartbeat tracking task** **1) Performance accuracy** Participants were asked to count their heartbeats over 6 epochs (25s to 50s) and performance accuracy derived from proximity of reported heartbeat number with the actual number of heartbeats recorded veridically. However, there were no patient vs control differences in performance accuracy on this task. Performance accuracy across patients did not correlate with affective symptoms and, despite visibly lower accuracy in the group of patients with clinical diagnosis of schizophrenia, the overarching test for between patient group differences did not reach criterion for significance. **2) Confidence** After each epoch of counting, the participant reported the number of perceived heartbeats and then gave a judgment of how confident they felt about their response accuracy. Patients were significantly less confident than controls in their performance (patients vs controls; VAS mean ± SD: 4.2 ± 2.6 vs 5.4 ± 1.8, F(1)=8.82, *P*<.005). This effect remained significant with age, sex, and BMI covariates (patients vs controls; VAS mean ± SDSD 4.2 ± 2.6 vs 5.5 ± 1.8, F(1)=10.3, P=0.001). Depression score correlated negatively with confidence in performance on the heartbeat tracking task (R=-0.11 P<005). Trait anxiety similarly correlated negatively with confidence on the heartbeat tracking task (R=-14, *P*<.01). However, between diagnostic patient groups, differences in mean confidence did not reach criterion significance. **3) Insight: accuracy confidence correspondence** We correlated scores for performance accuracy with confidence measures for each participant to derive a metric of metacognitive insight i.e. how well their confidence matched their performance. In the comparison of controls versus patients, there were no significant differences in metacognitive awareness (insight) into interoceptive ability. Similarly, difference across patients did not reach significance and this insight measure did not predict diagnostic group membership or correlate with affective symptoms **B) Heartbeat discrimination task** **1) Performance accuracy** Over repeated trials, participants were asked to judge if a series of notes were in time with their heartbeats or out of phase Patients’ performance accuracy was significantly lower than controls (patients vs controls without covariates; mean ± SD: 0.52 ± 0.1 vs 0.57 ± 0.2, F(1)=4.8, *P*<.05; with age, sex and BMI covariates F(1)=4.7, *P*<.05). **2) Confidence** After each trail of heartbeat discrimination, the participant gave a judgment of how confident they felt about their response accuracy. Overall patients were significantly less confident in their performance compared to controls (patients vs controls without covariates; VAS mean ± SD: 4.2 ± 2.6 vs 5.4 ± 1.8, F(1)=8.82, *P*<.005; with age, sex and BMI covariates, F(1)=6.3, *P*<.02). We observed no significant correlation between confidence measures on this task and affective symptomatology **3) Insight: accuracy confidence correspondence** Using Receiver Operator Characteristic (ROC) curves we compared correlated scores for discrimination trial-by-trial performance accuracy with confidence measures for each participant to derive a metric of metacognitive insight i.e. how well their confidence matched their performance. In the comparison of controls versus patients, there were no significant differences in metacognitive awareness (insight) into interoceptive ability. Similarly, difference across patients did not reach significance and this insight measure did not predict diagnostic group membership or correlate with affective symptoms.

On the heartbeat discrimination task, patients again showed lower confidence than controls (patients vs controls; VAS: 5.0 ± 2.4 vs 5.9 ± 1.8 [F(1,332)=5.85, *P*=.02]; with covariates [F(1,317)=6.3, *P*=.01]). Also, patients’ performance accuracy was lower (patients vs controls 0.52 ± 0.1 vs 0.57 ± 0.2, [F(1,331)=4.8, *P*=.03]; with covariates [F(1,316)=4.8, *P*=.03]). Again, groups did not differ in metacognitive insight on this task (0.52 ± 0.14 vs 0.54 ± 0.14, [F(1,327)=4.8, *P*=.38].

Self-rated interoceptive sensibility (BPQ, awareness scale) did not distinguish patients from controls (0.05 ± 1.5 vs 0.06 ± 1.4 [F(1,230)=4.8, *P*=.64]). Similarly, there were no group differences in interoceptive trait prediction error (ITPE 112.2 ± 29 vs 114.5± 34 [F(1,323)=4.8, *P*=.89]). Thus, patients differed from controls in the impact of cardiac signals on emotional processing, and showed reduced confidence and sensitivity in judging their own cardiac sensations. These effects were not primarily attributable to differences between controls and patients in physiology, subjective awareness or metacognitive representation of internal bodily state.

### Correlations with affective symptoms

In patients (controls excluded), depressive score (BDI score) correlated with state and trait anxiety (STAI-Y1 *R*(306)=0.54, *P*<.001; STAI-Y2 *R*(306)=0.74, *P*<.001). However, we found no initial correlation between depression score and heart rate, HRV, or interoceptive measures, except subjective interoceptive sensibility (BPQ-awareness, *R*(282)=0.34, *P*<.001) and ITPE (*R*(199)=0.22 *P*=.002). When we controlled for baseline covariates, depression score remained positively correlated with anxiety, subjective interoceptive sensibility (BPQ –awareness *R*(152)=0.31, *P*<.001), and ITPE (*R*(152)=0.24, *P*<.002). Depression score also then showed a weak negative correlation with HRV (RMSSD *R*(152)=-1.7, *P*=.04). Thus, across all patients, depressive symptoms were associated with anxiety, heightened subjective sensitivity to bodily sensations and (weakly) withdrawal of parasympathetic cardiac tone.

Anxiety symptoms correlated with depression score but did not correlate with heart rate (STAI Y1 *R*(298)=0.11, *P*=.07; STAI-Y2 *R*(298)=0.02, *P*=.073), nor HRV (STAI Y1 *R*(298)=0.03, *P*=.63; STAI-Y2 *R*(298)=0.05, *P*=.37). However, trait anxiety correlated with cardiac timing effects on fear processing (*R*(276)=0.13 *P*=.02), and metacognitive insight on heartbeat discrimination (*R*(284)=0.18, *P*=.002). State and trait anxiety also correlated with subjective interoceptive sensibility (BPQ-awareness; STAI-Y1 *R*(282)=0.33, *P*<.001; STAI-Y2 *R*(282)=0.39, *P*<.001) and ITPE (STAI-Y1 *R*(282)=0.23, *P*<.001; STAI-Y2 *R*(282)=0.22, *P*<.05). When we controlled for baseline covariates, trait anxiety remained correlated with cardiac timing effects on fear processing (*R*(152)=0.17, *P*=.04), and state and trait anxiety remained correlated with interoceptive sensibility (BPQ-awareness; STAI-Y1 *R*(152)=0.26, *P*=.001; STAI-Y2 *R*(152)=0.33, *P*<.05) and ITPE (STAI-Y1 *R*(152)=0.16, *P*=.05; STAI-Y2 *R*(152)=0.17, *P*=.04). Thus, across patients, anxiety symptoms were associated with increased subjective sensitivity to bodily sensations. However, transdiagnostic links between affective symptoms and either physiology or cardiac effects on emotional processing were weak, and we found no overarching association with heartbeat detection ability.

### Medication effects

We tested for general effects of antipsychotic and antidepressant medication. Patients taking only other medications (including Benzodiazepines, Pregabalin, Valproate, and Lithium) were excluded due to small group sizes, N<10. Significant findings were limited to: (1) Marginally higher heart rate in people on antipsychotics, an effect lost when controlling for baseline covariates (antipsychotic vs no antipsychotic; bpm 73.8 ± 11.5 vs 72.7 ± 11.3 [F(1,276)=4.76, *P*=.03]; with covariates [F(1,264)=0.39, *P*=.53]). (2) Significantly lower HRV in patients on antipsychotics (RMSSD ms: 42.1 ± 31 vs 58.5 ± 47 [F(1,268)=9.9, *P*=.002], with covariates F(1,276)=4.8, *P*=.03). (3) Increased subjective interoceptive sensibility with antidepressant use (antidepressant vs no antidepressant; BPQ-awareness scale: 116.4 ± 29.4 vs 108.8 ± 27.6 [F(1,269)=4.82, *P*=.03]; with covariates [F(1,258)=4.74, *P*=.03]). Thus, there was reduced HRV in patients taking antipsychotic medication. A weaker association between antidepressant medication and subjective interoception was consistent with an association with affective symptomatology.

### Differences between patient diagnostic groups on cardiac and interoceptive measures

We tested for differences in physiological and interoceptive measures between groups of patients categorised according to recorded clinical diagnoses. Diagnostic groups included depression (N=58), ‘anxiety disorder’ (N=28), mixed anxiety /depression (N=41), bipolar disorder (N=53), emotionally unstable personality disorder (N=21), ‘schizoaffective disorder’ (N=20) and schizophrenia (N=18) (See Methods and Table 1).

No effect of group emerged for heart rate ([F(6,241)=0.36, *P*=.92]; with covariates [F(6,232)=0.83, *P*=.55]; but there was an effect on HRV [(F(6,234)=3.3, *P*=.004]; with covariates [F(6,226)=2.4, *P*=.03]). There was a differential cardiac timing effect on emotional face processing ([F(6,234)=3.1, *P*=.006]; with covariates [F(6,226)=2.8, *P*=.01]) and a trend for this effect on fear alone ([F(6,238)=2.1, *P*=.06]). Objective performance accuracy, subjective confidence in heartbeat perception, and metacognitive interoceptive awareness of heartbeat, did not discriminate clinical groups (Fig. 3). However, groups differed in subjective interoceptive sensibility (BPQ-awareness scale [F(6,236)=2.4, *P*=.03]; with covariates [F(6,228)=1.8, *P*=.10] trend) (Fig 3).

Decreased HRV characterised patients with diagnoses of bipolar disorder, emotional unstable personality disorder, schizoaffective disorder, and schizophrenia, relative to anxiety and depression groups (and controls). Interestingly, for cardiac effects on emotional processing, the normative modulation of fear and neutral processing were reversed in patients with a primary clinical diagnosis of schizophrenia (Fig. 2). This reversed effect was not observed for patients in the ‘schizoaffective’ group (exploratory analysis schizophrenia vs schizoaffective; interaction [F(1,40)=6.9, *P*=.01][; with covariates F(1,36)=7.9, *P*=.008]; fear [F(1,40)=3.15, *P*=.08]; with covariates [F(1,36)=6.0, *P*=.02]; neutral [F(1,40)=5.7, *P*=.02]; with covariates F(1,36)=4.3, *P*=.045). This group difference was not accounted for by medication (group x antipsychotic effect, interaction [F(1,30)=0.01, *P*=.94]) nor by illness duration, retrieved for schizophrenia and schizoaffective patient records (i.e. cardiac timing effect remained significant when illness duration was included as confounding covariate (all covariates: [interaction F(1,34)=6.0, *P*=.02]). Higher scores for depression (BDI), anxiety (STAI) in the schizoaffective group were subthreshold for significance.

## DISCUSSION

In a representative patients using mental health services in the UK, we characterized interoception, i.e. processing and representation of internal bodily physiology^1-5^. We predicted a transdiagnostic relationship between interoception and psychopathology^1-5,9,13,15,17,18^, especially anxiety symptoms, previously linked to differences in physiology (heart rate and HRV)^29,46^, cardiac effects on emotional processing^33,34^, heartbeat detection accuracy^21,22,24^, self-reported bodily sensibility^47^, and discrepancies between objective and subjective measures of interoception^20,25^.

Correspondingly, we found that patients differed from controls in physiology (HRV), cardiac effects on emotional processing, subjective ratings of interoceptive performance accuracy (heartbeat discrimination), and subjective interoceptive confidence. Across patients, subjective interoception paralleled affective symptoms: Lower interoceptive confidence alongside higher subjective interoceptive sensibility (self-rated trait interoceptive ‘beliefs’) and interoceptive trait prediction error (ITPE; divergence of subjective interoceptive sensibility from objective interoceptive accuracy^25^), correlated with affective symptoms. However, such symptoms showed limited transdiagnostic association with physiology, despite established coupling of perseverative cognition (worry and rumination) to lower HRV^48^. Reduced HRV correlated weakly (after covariate correction) with depressive symptoms. Moreover, HRV was observed to be lower in diagnoses other than depression and/or anxiety disorder (see below)^46^.

Our hypothesis that cardiac effects on emotional processing would predict anxiety symptoms, was partly supported: Across patients, enhancement of fear processing by systole (i.e. arterial baroreceptor signals) correlated with trait anxiety. Moreover, patients with a unitary diagnosis of anxiety disorder (i.e. generalised anxiety disorder or panic) showed the strongest systolic enhancement of fear processing^34^. Overall however, our findings endorse a more general association between subjective aspects of interoception and affective symptoms.

Our data also demonstrates differences in interoception between psychiatric diagnostic categories. First, our findings extend previous reports of markedly reduced HRV in patients with emotionally unstable (borderline) personality disorder^49^, bipolar disorder^50^ or schizophrenia/psychosis^51,52^. Speculatively, this may reflect vulnerability to dissociative states. Cardiac effects on emotional processing also differentiated patient groups: While patients with an anxiety diagnosis showed strongest systolic enhancement of fear, patients with clinical diagnosis of schizophrenia stood out from all other groups in showing an opposite cardiac effect on emotional processing. This difference even contrasted with the normative response observed in patients with schizoaffective disorder/unspecified psychosis, whose symptomatology and (antipsychotic) medication overlaps with schizophrenia. Trends toward faster mean heart rate and worse heartbeat tracking accuracy suggest more pervasive interoceptive deficits in schizophrenia. Schizophrenic and schizoaffective groups were also differentiated by the schizoaffective patients significantly under-reporting subjective sensitivity^20^ to bodily sensations. The clinical distinction between schizophrenia and schizoaffective disorder is rarely examined in research studies, favouring instead a broader diagnosis of psychosis. Although psychotic phenomena suggest disordered self-representation^13-19^, our study’s focus on transdiagnostic relationships between interoception and affective symptoms meant that we did not quantify psychotic and dissociative symptoms. However antipsychotic medication, and illness duration, did not provide a compelling account for interoceptive differences in schizophrenia. Thus, our exploratory findings motivate further research to characterise schizoaffective and schizophrenic symptoms with attention to interoceptive profiles^17^.

Heartbeat detection tasks seek to quantify stable individual differences in sensitivity to cardiac sensations. Typically the heartbeat counting task gives a spread of accuracy scores, while the harder heartbeat discrimination task produces a more binomial distribution (i.e. at chance, or above chance). Nevertheless, these tasks have recognised psychometric limitations^28,53^: Actual heart rate, knowledge of one’s average heart rate, and the ability to estimate time, can influence performance accuracy particularly on heartbeat tracking. The perceived signals themselves may be ‘quasi-interoceptive’, i.e. somatosensory correlates of internal physiology^53^. These factors contaminate objective measurement of interoceptive sensitivity with subjective beliefs and predictions about what should be felt (sensibility^20^). Notwithstanding, heartbeat detection tasks remain relevant to inferences about how bodily-sensations influence emotional states. In non-clinical populations, heartbeat detection ability is frequently associated with anxiety symptoms, yet attenuated by depressive symptoms^23,24^. In schizophrenic patients, reduced heartbeat detection accuracy reportedly correlates with positive psychotic symptoms^17^. Within our study, patients performed worse than controls on the heartbeat discrimination task, though among patient groups, performance accuracy was broadly equivalent (schizophrenic patients tended to perform worse). While interoceptive methods can be further optimised for patient stratification, we demonstrate reliable implementation of heartbeat detection tasks within clinical settings.

In psychiatry, interoception is a treatment target. Medications influence interoceptive processes; e.g. peripheral cardiovascular arousal is suppressed by beta-blockers, while monoaminergic drugs (from stimulants in ADHD, to antidepressant / anxiolytic SNRIs) target central neuromodulatory pathways governing central arousal and descending autonomic control. Trials repurposing antihypertensive drugs, e.g. Losartan^55^, and research on interoceptive immune-brain and gut-brain signalling^3,10,11,54^ promise alternative treatment approaches. Non-pharmacological therapies also exploit interoceptive mechanisms. These include physical interventions, notably vagus nerve stimulation^56^, bio-behavioural therapies, (e.g. autonomic biofeedback training)^57^, and integrative interventions (e.g. mindfulness and yoga)^58^, whose therapeutic utility can be optimised with better mechanistic understanding of interoceptive actions.

This study follows a timely call for interoceptive processes to be considered foundational to psychiatric disorders^1^. We show the feasibility of a multilevel characterization of interoception in patients spanning diagnostic categories. Our findings reveal transdiagnostic interoceptive deficits linked to mood symptoms, and suggest interoceptive measures can differentiate patients by diagnosis, notably potentially selective abnormalities in schizophrenic patients that merit further investigation. Interoception thereby offers emerging targets for therapeutic intervention in psychiatric disorders.

## Data Availability

Anonymised data will be made available after peer-review publication

## Acknowledgements

The study was funded by an Advanced Grant from the European Research Council to HDC: *Cardiac control of Fear in the Brain CCFIB 324150*. Recruitment and data collection to the study was facilitated by Sussex Partnership NHS Foundation Trust Research and Development Office and the input of 3 Clinical Research Coordinators.

## Notes

### Competing Interest Statement

The authors have declared no competing interest.

### Clinical Trial

ISRCTN13588109

### Author Declarations

All relevant ethical guidelines have been followed and any necessary IRB and/or ethics committee approvals have been obtained.

Any clinical trials involved have been registered with an ICMJE-approved registry such as ClinicalTrials.gov and the trial ID is included in the manuscript.

